# Intestinal microbiota changes due to *Giardia intestinalis* infections in a longitudinal Ecuadorian birth cohort and impact on cobalamin biosynthesis

**DOI:** 10.1101/2024.10.15.24315510

**Authors:** Ashish Damania, Andrea Arévalo-Cortés, Andrea Lopez, Victor Seco-Hidalgo, Diana Garcia-Ramon, Emilie Lefoulon, Courtney Long, Evan Drake, Barton Slatko, Philip J Cooper, Rojelio Mejia

**Affiliations:** Platform for Innovative Microbiome and Translational Research, Department of Genomic Medicine, The University of Texas MD Anderson Cancer Center, Houston, Texas; School of Medicine, Universidad Internacional del Ecuador, Quito, Ecuador; Faculty of Human Health, Universidad Nacional de Loja, Loja, Ecuador; Institute of Infection and Immunity, St George’s University of London, London, UK; Genome Biology Division, New England Biolabs, Inc, Ipswich, MA, USA; Department of Pediatrics – Tropical Medicine, Baylor College of Medicine, Houston, Texas

**Keywords:** microbiome, cobalamin, vitamin B12, *Giardia intestinalis*

## Abstract

**Background:** *Giardia intestinalis* is a protozoal parasite infecting the gastrointestinal tract worldwide. Chronic infections/reinfections are common, with adverse nutritional consequences for critical growth during the first five years of life. Vitamin B12 (cobalamin) is absorbed primarily in the ileum of the small intestine, where *Giardia* trophozoites attach and replicate. Bacteria activate bioavailable vitamin B12, which is essential for human DNA synthesis and development. A disturbance in cobalamin biosynthesis caused by giardiasis may contribute to impairment of childhood development and growth.

**Methodology/Principal Findings:** We performed a longitudinal analysis on 61 Ecuadorian children using multi-parallel real-time quantitative PCR and whole genome sequencing. Children had increased *Giardia* DNA burden from ages 3 to 5 (p = 0.0012) and 7.58 times more frequent *Giardia* infections (1.31 to 34.33, P = 0.0176). There was an increased alpha diversity/ *Giardia* fg/µl in three-year-olds compared to age-matched non-infected (30.20 vs 4.37, p = 0.050), but decreased alpha diversity/ *Giardia* fg/µl in five-year-olds compared to age-matched non-infected children (0.21 vs 4.31, p = 0.021). Alpha diversity/ *Giardia* fg/µl was also decreased in samples collected longitudinally from the same children at five compared to 3 years (p = 0.031). Cobalt transport protein (*Cbi*N) (FDR < 0.003) and IPR011822 (Cobalamin-dependent methionine synthase) sequences were decreased in infected children (p < 0.002) and among those with greatest *Giardia* burdens (p < 0.001).

**Conclusion/Significance:** *Giardia intestinalis* infection may affect bacterial diversity in the ileum where vitamin B12 is activated, as suggested by a reduced proportion of Cobalt transport protein component (C*bi*N) gene sequences in the gut microbiome of infected children. These findings are potentially important to our understanding of how *Giardia* infections may affect childhood growth.

## Background

Giardiasis is a common parasitic disease in humans in low-income settings. It is caused by the protozoan *Giardia intestinalis* (syn. *G. duodenalis* or *G. lamblia*) that infects humans through the ingestion of cysts present in a fecally-contaminated environment. Trophozoites, released in the stomach, adhere to and replicate in the small intestine, while in the jejunum, trophozoites transform back into cysts that are excreted in feces. Infections cause clinical manifestations ranging from an asymptomatic state to acute or chronic diarrhea, malabsorption, abdominal pain, vomiting, and nausea. Differences in clinical presentation may vary according to the infecting *Giardia* assemblage, infectious dose, age and nutritional status of the host, and specific effects of *Giardia* on gut microbiome composition (1,2) (3–6).

*G. intestinalis* is a common infection in children living in poorly hygienic conditions, and chronic infections impair growth and cognition (2). The prevalence of giardiasis among children in some of Ecuador’s peri-urban and rural regions has been estimated between 20% and 31.5% (7–9). In contrast, in samples of children from Brazil and Colombia, prevalence was 44% and 60%, respectively (10,11).

The vital role of maternal and early-life nutrition in physical and neural development (12) has been well described. Vitamin B12, or cobalamin, is a critical micronutrient exclusively biosynthesized by specific microbes in the gastrointestinal tract. Vitamin B12 is required during infancy and childhood to develop higher brain functions such as language and cognition. A deficiency can result in megaloblastic anemia and neurological complications such as sensory and motor disturbances and cognitive declines (13,14) (15). Vitamin B12 deficiency has been linked to decreased intrinsic factors, bacterial overgrowth, and parasitic infections, including *Giardia* (13,16). Cobalamin deficiency can also cause bacteria to shift elsewhere in the gastrointestinal tract and could disrupt microbial interactions (17).

Few studies have examined the interactions between the gut microbiome and vitamin B12. Degnan et al. (2014) described how most of the known human gut microbial species required B12 but lacked the genes necessary to synthesize B12 *de novo* - less than 25% of intestinal bacterial species can synthesize vitamin B12(18). Vitamin B12 does not appear to affect the intestinal microbiome: an analysis of infants aged 4-6 months did not show alterations in the intestinal microbiota following vitamin B12 supplementation of vitamin B12 deficient infants (19). In contrast, vitamin B12 supplementation in patients with homocystinuria did not affect the alpha or beta diversity of the gut microbiome (20).

*Giardia* infections could affect vitamin B12 through its effects on the intestinal microbiome. Mejia et al. (2020) evaluated the potential impact of giardiasis on the interaction between the gut microbiome and vitamin B12, showing that *G. intestinalis* infected children had a higher abundance of the genus *Prevotella,* and reduced presence of *Cbi*M, a gene involved in vitamin B12 biosynthesis, compared to uninfected children (21). Other protozoas, including *Giardia,* also show shifts in microbial communities(22).

In the present study, we hypothesized that *Giardia* infections alter the gut microbiome, causing changes in bacterial species that activate vitamin B12 and may deplete bioavailable vitamin B12, contributing to growth impairment in young children. To test this hypothesis, we did a taxonomic and metagenomic analysis of the bacterial microbiome from fecal samples collected longitudinally from Ecuadorian children at 3 and 5 years of age who were infected (or not) with *G. intestinalis* as determined using multi-parallel real-time quantitative PCR (qPCR). Our analyses of the bacterial microbiome of the gut focused on the diversity of bacteria and their vitamin B12 activating sequences.

## Methods

### Sample selection and study procedures

For the present study, we used a nested sample of children that were randomly selected from a birth cohort of 2,404 newborns followed up from birth to 8 years of age and living in the rural District of Quininde, Esmeraldas Province, in tropical coastal Ecuador (23). For the present study, we selected 61 stool samples from children collected at 3 and 5 years of age with 1) a known positive or negative result for *G. intestinalis* DNA using qPCR and 2) who were negative for the presence of DNA from other gastrointestinal parasites (*Ascaris lumbricoides, Ancylostoma duodenale, Necator americanus, Strongyloides stercoralis, Trichuris trichiura, Cryptosporidium* species, and *Entamoeba histolytica*) (9). None of the children had received antibiotics within three months of stool samples being collected. After the study sample collection, all children with positive gastrointestinal parasite results were treated with appropriate anti-parasitic medication. Paired stool samples at 3 and 5 years were available for 27 children, while unpaired samples from 7 children were available at 3 or 5 years of age.

Data on sociodemographic, individual, and household factors were collected by maternal questionnaire around the time of birth of the child. A trained member of the study team administered the questionnaire.

### Ethics statement

Informed written consent was obtained from a parent or guardian of each child. The study protocols were approved by the Bioethics Committee of the Universidad Internacional De Ecuador, Quito, Ecuador. From the parent study, the results of stool microscopy were given to the parent/guardian and anti-parasite treatment was provided where appropriate. The study was also approved by the Baylor College of Medicine, institutional review board H-33219.

### Parasite sample preparation

Stool samples were stored in ice and air-tight containers immediately after collection for later DNA extraction using MP FastDNA Spin Kits for Soil (MP Biomedicals, Irvine, CA). DNA from samples stored at -20° C and processed by multi-parallel qPCR (*G. intestinalis, A. lumbricoides, A. duodenale, N. americanus, S. stercoralis, T. trichiura, Cryptosporidium* species*, and E. histolytica)* as described (9,24).

### Microbiome sequencing

Microbiome sequencing was performed at New England Biolabs, Inc. Preparation included removing methylated DNA with NEBNext ^®^ Microbiome enrichment kit with no size selection. All samples used one µg of DNA with eight cycles of PCR enrichment (NEBNext Ultra DNA Library Prep Kit for Illumina, Version 5.1, 9/17). Illumina NextSeq (Illumina, San Diego, CA) or Illumina MiSeq (Illumina, San Diego, CA) was used to produce 151bp reads (single and paired-end); however, only NextSeq sequencer reads were retained for downstream analysis.

### Bioinformatics analysis

Paired ends were interleaved using the BBMap program (version 38.73) to convert all fastq reads to single-end reads for downstream processing (25). All single-end fastq reads were filtered using a fastp program set at a minimum Phred score of 15 and a minimum length set at 50bp (26). MultiQC was used to visualize and summarize the filtered fastq reads (27). The NR database was downloaded in fasta format from ftp://ftp.ncbi.nlm.nih.gov/blast/db/FASTA/ on 19^th^ December 2019 as the reference database (28). Filtered fastq reads were processed using Diamond (Version v0.9.29) with blastx mode at the default setting and maximum target sequences set to one (29). Output from Diamond was processed to extract taxonomy and InterPro protein database results using the Megan (community edition version 6.20.17) program in STAMP format (30–32).

### Taxonomic and metagenomic analysis

The resultant taxonomy text file was imported into Phyloseq for calculating Shannon alpha-diversity (species richness) and beta-diversity (species variability) (33). The “dplyr” package was used for data transformation within the R programming language environment (34,35).

Taxonomic data were filtered to remove reads less than 500 reads in 20% (12 of 61 samples) of the total samples. Any reads from phylum chordate were removed to filter for possible human reads. Batch effects were investigated using an ordinate function from the Phyloseq package with the “bray” distance method and Principal Coordinate Analysis (PCoA). The InterPro database was searched for using the keyword "cobalamin" to manually extract all vitamin B12 biosynthesis and transporter genes using InterPro entry ID and description, which resulted in 142 proteins^[1]^.

The taxonomic and InterPro results from individual samples were exported in tab-delimited file format and subsequently merged using the "merge_metaphlan_table.py" Python script, which was included with metaphlan software version 2.2.0 (36). Vegan package was used to determine beta-diversity metrics, including PERMANOVA and beta-dispersion among the *Giardia* quartile (zero to three), reflecting the intensity of *Giardia* DNA found via qPCR, *Giardia* infection status and anthropometric scores (37). MaAsLin2 was used to test for differentially abundant taxa with minimum prevalence of 25% and minimum abundance of 100 reads (38). *Giardia* parasite burden was measured in 4 ordinal groups that included uninfected children and the 3 of 4 quartiles of increasing concentrations of *Giardia* DNA (1^st^ (0.01 – 0.15), 2^nd^ (0.17-24.50) and 3^rd^ (118.58 – 29946.58). Differential analysis for genes from metagenomic data was performed using the corncob package (39) with “differentialTest” function. Statistically significant results were further run through “bbdml” function to obtain an effect size estimate using maximum likelihood. Lastly, “plot_bbdml” function on the bbdml output with 10,000 bootstrap simulations was used to obtain a 95% confidence interval of individual relative abundance values.

### Statistical analysis

The Kruskal-Wallis test was used to compare three or more independent groups. Pairwise Wilcox tests were used as post-hoc tests with a false discovery rate as a p-value correction method. Statistical significance was set at p < 0.05, and the false-discovery rate was set at 0.05 for all tests. Linear mixed effect models were conducted using the lmerTest R package (40). Graphs were created using Prism Version 10.3.0 (Graphpad Software, Boston, MA) and the R ggplot2 package (41).

## Results

### Comparing Giardia DNA burden and assemblage among infected children

There was a significant increase in the *Giardia* DNA burden at five compared to three years among samples from all children with evidence of infection at both ages (p=0.0012) **(Figure 1A)**, and this was also seen in paired samples (p=0.031) **(Figure 1B)**.

**Figure 1:**
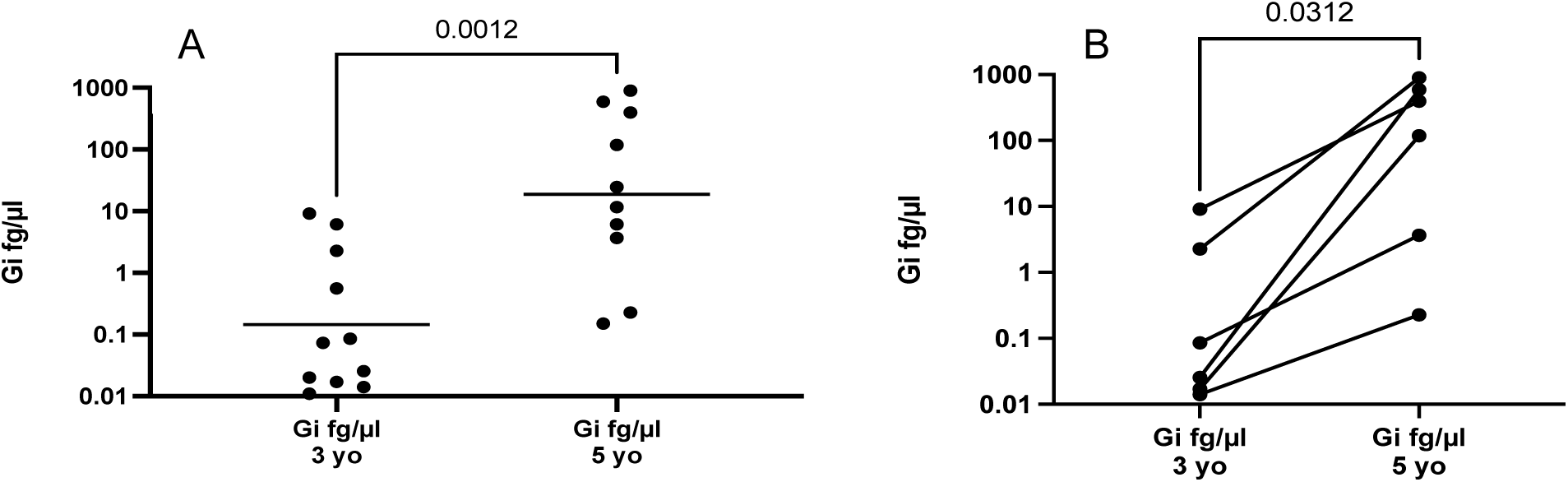
A. Higher levels of *Giardia* DNA in 5-year-old infected children compared to three-year-old infected children (0.145 vs 18.76 fg/µl, P = 0.0012). B. In individually *Giardia-infected* children, there was a significant increase in *Giardia* DNA burden in paired samples (P = 0.0312).

If a 3-year-old was infected with *Giardia,* there were 7.58 greater odds (95% CI 1.31-34.33, P = 0.0176) of having giardiasis at five years old **(Figure 2)**. Using A and B assemblage sequences, we detected 45.9% assemblage A, 49.2% assemblage B, and 82.1% with both A and B assemblages. Because of the large percentage of mixed assemblages in this cohort, assemblages were not considered for the analysis.

**Figure 2.**
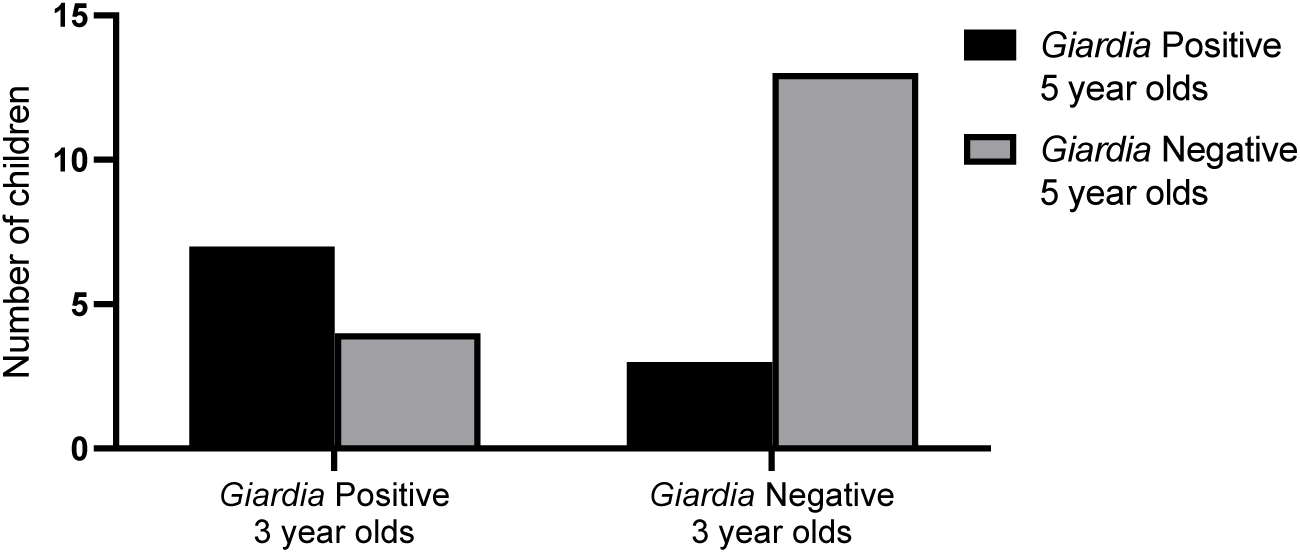
Children infected with *Giardia* at three years had a 7.58 greater odds of being infected at five years (1.31 to 34.33, P = 0.0176).

**Figure 3.**
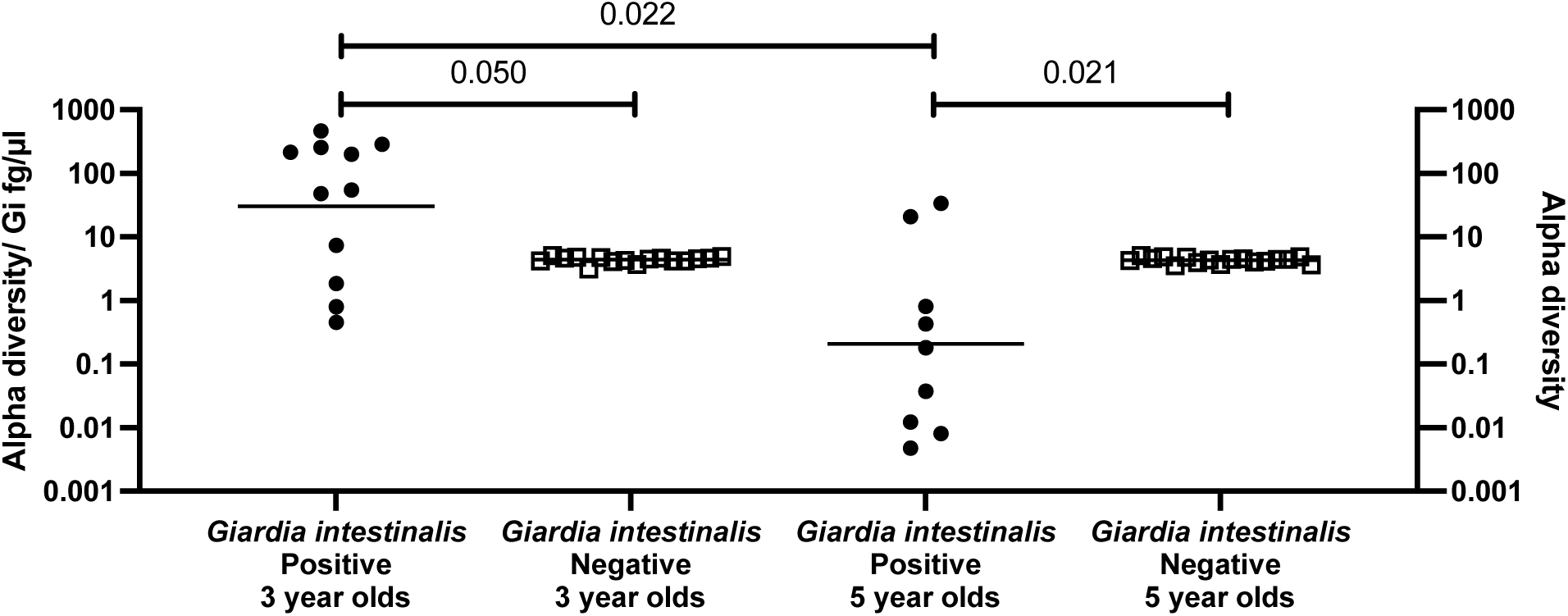
Alpha diversity normalized to concentration (fg/µl) of *Giardia intestinalis* DNA in infected children aged three-year-olds was higher than in non-infected three-year-olds (30.20 vs 4.37, p = 0.050). The opposite was seen in five-year-olds infected and non-infected (0.21 vs 4.31, p = 0.021). There was also a decrease in alpha diversity/ *Gi* fg/µl in the five-year-olds and an overall difference between all cohorts (p = 0.0053). For children without *Giardia,* their results are on the Y2 axis and are labeled Alpha Diversity.

### Alpha diversity of infected and non-infected children

Alpha diversity, measuring gut microbiome species diversity using the Shannon index, did not differ significantly by *Giardia* parasite burden between these two groups; there were no significant differences relative to *Giardia* DNA abundance **(Supplemental Figure 1)** or between age or sex **(Supplemental Figure 2)**.

To incorporate the impact of *Giardia* DNA burden on bacterial alpha diversity, a ratio of alpha diversity was divided by *Giardia* DNA fg/µl was calculated for each sample. There was some evidence of increased alpha diversity/ *Giardia* fg/µl in three-year-olds compared to age-matched non-infected (30.20 vs. 4.37, p = 0.050), but decreased alpha diversity/ *Giardia* fg/µl in five-year-olds compared to age-matched non-infected children (0.21 vs 4.31, p = 0.021).

Individual children that were *Giardia* positive at both time points at three and five years showed a decrease in the ratio of alpha diversity relative to *Giardia* DNA concentration fg/µl (p = 0.022). In the presence of *Giardia* infection, alpha diversity was greater in relation to *Giardia* DNA (fg/µl) in three compared to five-year-olds and also compared to *Giardia*-negative children (Kruskal-

In paired samples, there was a similar decrease in Alpha diversity/ *Gi* fg/µl in the five-year-olds (p = 0.031) with no differences in the other paired groups **(Figure 4)**.

**Figure 4.**
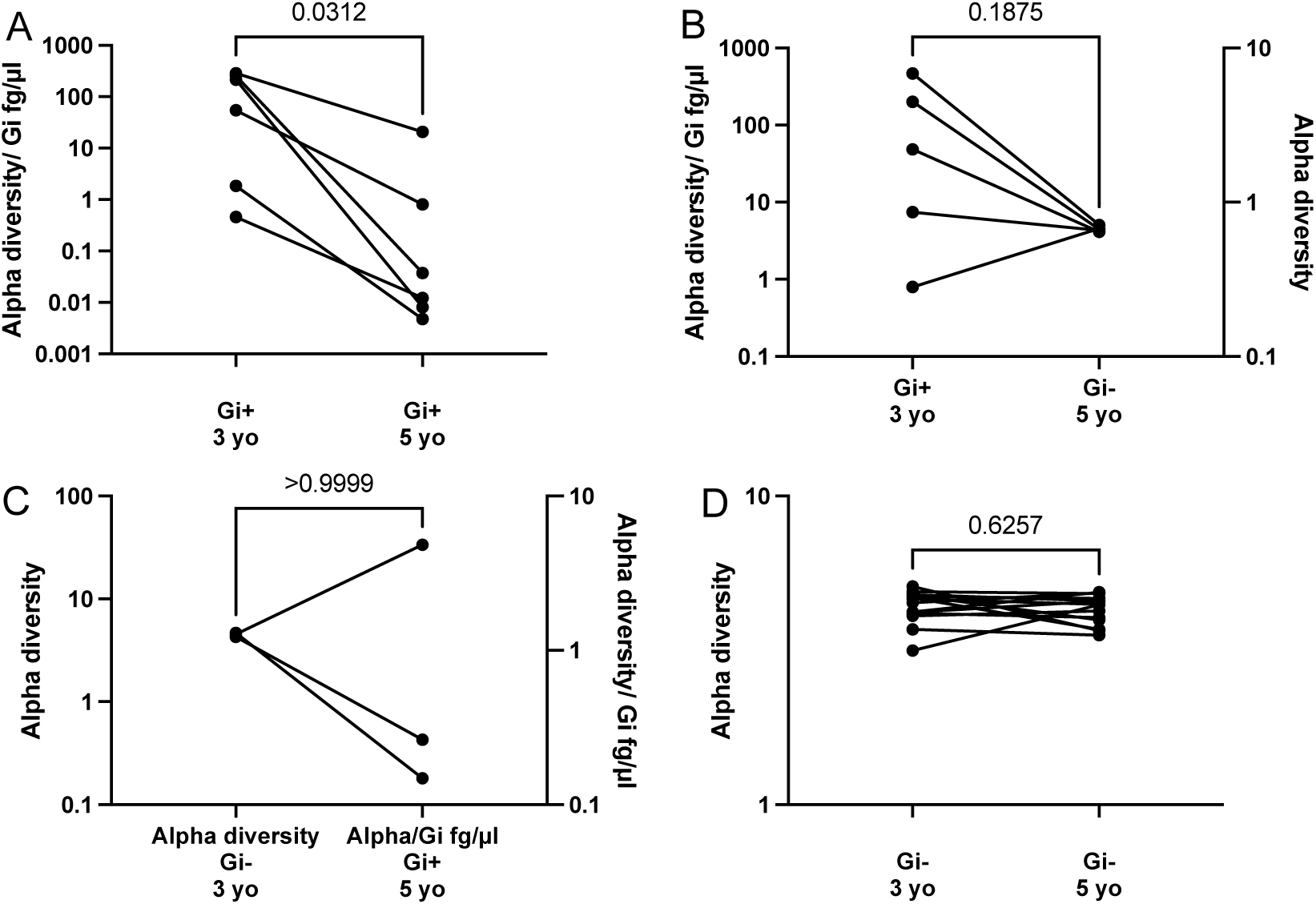
Paired results of Alpha diversity normalized to *Giardia* DNA concentrations (fg/µl) between infected and non-infected children. There was a decrease in Alpha diversity/ *Giardia* fg/µl in five-year-old children compared to three-year-olds (p = 0.031).

### Beta Diversity in Giardia infected and non-infected children

For three-year-old children, there was no difference in beta diversity between *Giardia, which was* positive and negative (R^2^ = 0.032, p = 0.401). For five-year-old children, there was no difference in Beta diversity between *Giardia* positive and negative (R^2^ = 0.0262, p = 0.837) **(Supplemental Figures 3)**. Exploration of independent effects of *Giardia* infection status or *Giardia* quartile, age, and sex on distance matrix calculated using PERMANOVA did not reveal any statistically significant terms to explain variance **(Supplemental Table 1)**. Variance in beta diversity between groups did not differ by *Giardia* infection status or across ordinal groups of parasite burden. Beta dispersion plots, ANOVA, and permutation tests with 10,000 permutations confirmed no significant difference in variance by *Giardia* infection quartiles for three-year-olds (p = 0.842) or five-year-olds (p = 0.796) **(Supplemental Figure 2)**.

### Differential Taxonomic differences

Differential abundance analysis was conducted to see possible changes at the species and genus level with respect to infection status, the latter allowing comparisons with other 16S rRNA gene sequencing studies. MaAsLin2 analysis (differential test function) was done to identify OTUs that were differentially abundant relative to *Giardia* infection. Species were differentially abundant and positively enriched in uninfected compared to infected children in 3-year-olds **(Figure 5A)** and 5-year-olds **(Figure 5B)**. All genus and species OTUs shown have FDR < 0.05. At the species level, we saw bacteria belonging to the genera *Veillonella, Clostridium*, and *Enterobacter* more abundant in infected children. Uninfected children had abundant *Ruminococcus, Bacteroides,* and *Eubacterium* species compared to *Giardia-infected* children of the same age **(Figure 5)**

**Figure 5:**
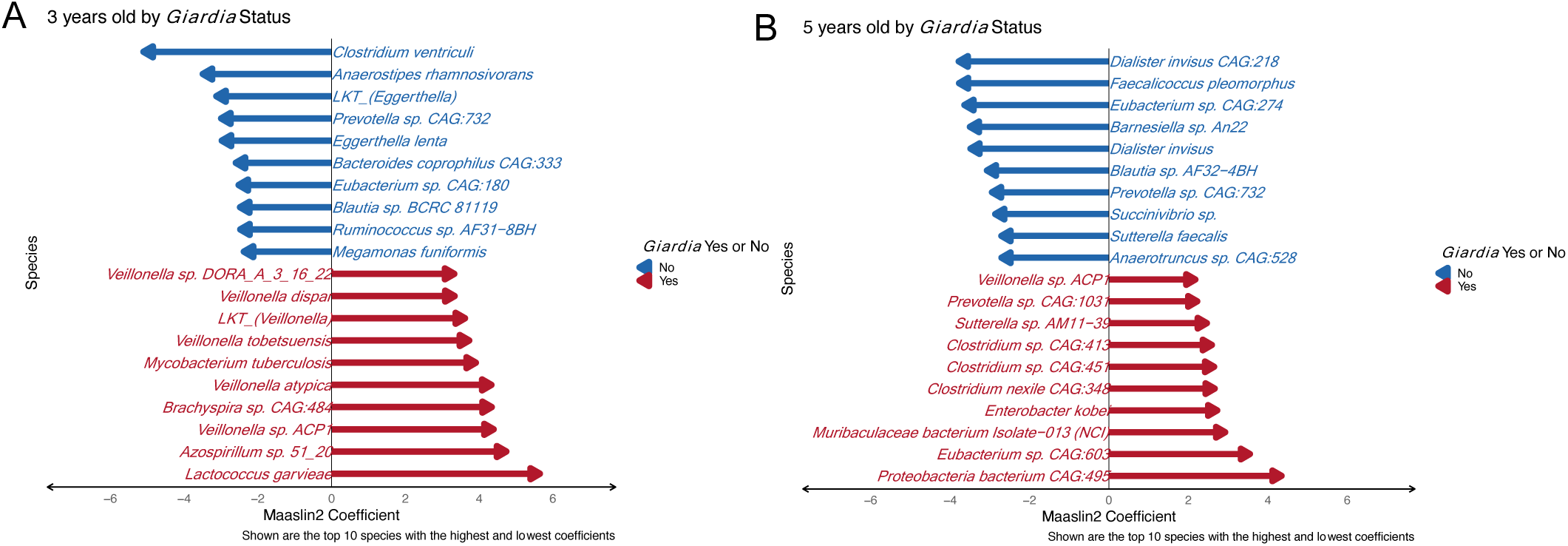
Top differentially abundant OTUs at species level comparison of *Giardia* infected versus uninfected children controlling for three-year-olds (A) and five-year-olds (B).

A comparison of paired samples infected with *Giardia* DNA with uninfected children showed 33 OTUs at genus and 132 at species levels to be differentially abundant. Differentially abundant taxa between these groups included the genera *Bacteroides sp. Ruminococcus sp.* and *Mycoplasma sp.* were also observed among the top 20 OTUs at the species level **(Figure 6A)**. Children who were initially not infected at three years old and then positive at 5-years-old had several *Prevotella* species **(Figure 6B)**. Three-year-old infected children had more *Bacteroides sp., Paraprevotella sp.,* and *Acinetobacter sp*. compared to non-infected five-year-olds had more *Ruminococcus sp, Bifidobacterium longum,* and *Eubacterium sp*. **(Figure 6C)**. Children of both ages had fewer bacterial differences than the other cohorts **(Figure 6D)**.

**Figure 6.**
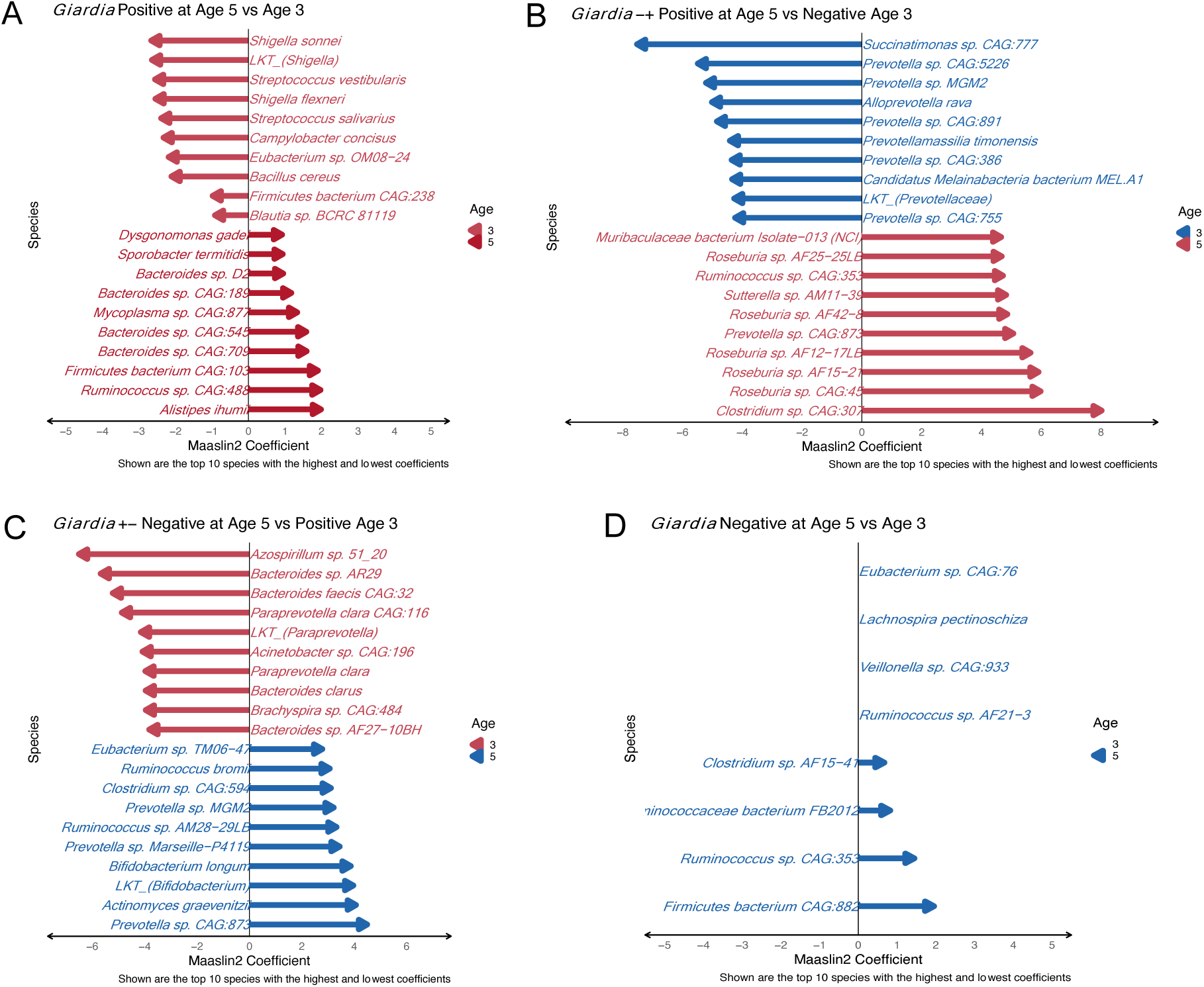
Top 20 differentially abundant OTUs at genus and species level for *Giardia* positives at both ages (A), the non-infected at three years old and then infected at five years old (B), and those infected at three years old and then non-infected at five years old (C). Only 8 OTUs were significantly different in abundance for those children not infected at both ages (D).

### Differential abundant cobalamin genes

Out of 143 vitamin B12 biosynthesis genes, 32 genes were found to be present among the 61 samples. When conducting a differential abundance test from the Corncob package by *Giardia* infection status using beta-binomial regression with sex and age as covariates, IPR003705 – Cobalt transport protein (*Cbi*N) was found to be significantly in low abundance in *Giardia-positive* children with FDR < 0.001. *CbiN* was further tested individually with sex and age as covariates. It was found to be less prominent in the *Giardia-positive* group with p = 0.002. The resultant graph shows the predicted confidence interval of the *Cbi*N gene in samples after 10,000 bootstrap simulations **(Figure 7A)**.

**Figure 7.**
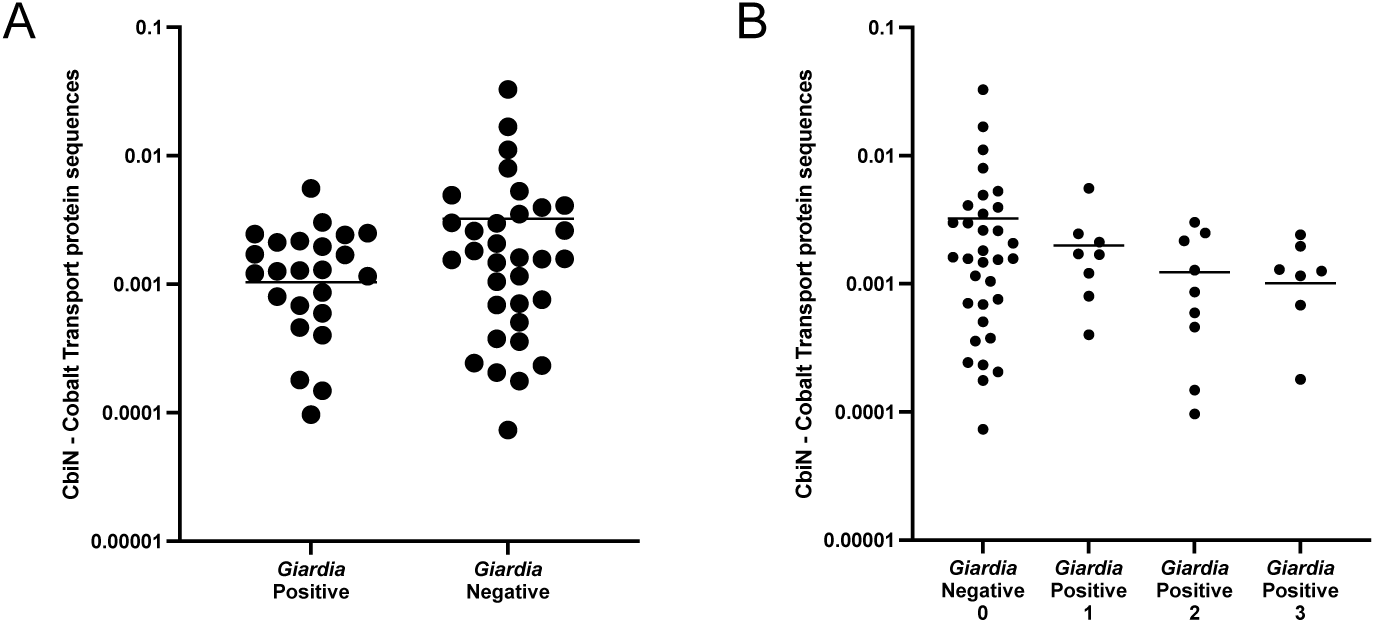
Relative abundance of the CbiN gene was decreased in children infected with *Giardia* compared to non-infected children (A). Relative abundance of CbiN gene among all the samples by *Giardia* concentration quartiles, Neg (0), and increasing concentrations (1, 2, 3). The prediction intervals are calculated using 10,000 bootstrap simulations.

Notably, the differential abundant test with *Giardia* quartile instead of *Giardia* infection status indicated genes coding for IPR003705 – Cobalt transport protein (*Cbi*N) (FDR < 0.003) and IPR011822 (Cobalamin-dependent methionine synthase) with FDR = 0.003 were found to be statistically significantly differentially abundant in the *Giardia* negative group vs. *Giardia* positive group, with sex and age as covariates.

Differential analysis of these genes was individually analyzed to get estimates by the *Giardia* quartile. For the *CbiN* gene, the remaining *Giardia* quartiles (2 and 3), as compared to the no *Giardia* quartile, had statistically significant negative abundances with p = 0.01 and p < 0.001, respectively **(Figure 7B)**. The predicted intervals from the differential abundance testing for *Cbi*N alone are noticeably higher in the *Giardia-positive* group.

## Discussion

*Giardia* infections are estimated to be present in about one-third of developing countries’ children and adult populations (42). Microbiome perturbation due to *Giardia* infection was minimal compared to *Giardia-negative* children, as there were no significant changes in the alpha-diversity and beta-diversity between the groups. Our results also show a non-significant increase in the alpha diversity with age. A study conducted on 417 Swedish children from newborn to five years of age found that the compositional variation was the least between three and five years (0.9%) (43). This indicates that microbiome changes between these time points are leveling out as they may be approaching an adult-like state. Such a decline in change may have flattened Giardia’s impact on diversity changes. Mice studies have shown an increase in Shannon alpha-diversity or richness in species following *Giardia* infection, while other studies have found no alterations (3,5,44). However, these studies were conducted with 16S rRNA sequencing and murine models. Microbiome studies in humans show that *Giardia* infection can reduce alpha diversity (6,21) or distinctive microbial profiles in individuals with asymptomatic versus symptomatic giardiasis(45). Toro-Londono et al. found significant changes in beta diversity in the *Giardia* group vs. the control group(46).

It is speculated that the amount of *Giardia*, or its burden, is related to disease progression, including changes in the intestinal microbiota(47). Therefore, we used the amount of *Giardia* DNA, in fg/µl, to modulate the microbial alpha diversity by mathematically dividing them and creating a ratio of alpha diversity/ *Giardia* fg/µl of DNA. This ratio explores how *Giardia’s* DNA burden affects microbial alpha diversity per infected child.

This study revealed a number of differentially abundant taxa, which may help understand the microbiome changes caused by the *Giardia* infection. It was not possible to distinguish the stool sample bacteria from the various intestinal regions. However, the small intestine has a higher oxygen level and less bacteria density than the colon due to the antimicrobial activity of bile acids (48). Samples with the highest *Giardia* infection showed abundant anaerobic species from *Clostridium* and *Veillonella* (**Figure 5**), which reside in the duodenum and cause biofilms in vitro (49,50). Biofilms can lead to pathogenesis, antimicrobial resistance, virulence, and restructure the microbiome (51). Interestingly, *V. atypica* from genus *Veillonella* is a lactate metabolizer in a low pH environment such as the small intestine and may negate some of the probiotic effects of lactic acid-producing bacteria *Lactobacillus* and *Bifidobacterium* on *Giardia* (52,53).

Cobalamin is absorbed by Intrinsic factor (IF) at the cobalamin complex at the receptors in the ileum. It is known that cobalamin absorption can be impacted by small intestine bacterial overgrowth (SIBO) due to competition with IF (54). Some microbes, such as *Bacteroides thetaiotamicron,* can remove cobalamin from IF with BtuG2 protein and compete directly with the human host (55). This study found bacteria containing *Btu*G homologs, such as *B. coprophilus,* with lower abundance in the highest *Giardia* quartile than the *Giardia* negative group.

Specific cobalamin biosynthesis sequences were decreased in the *Giardia-positive* groups, showing that the microbiome was deficient in cobalamin generation genes. Changes in microbiome diversity and the low presence of *Btu*G homologs bacteria may have been due to the effects of *Giardia* infection in these children. Corncob analysis limited to cobalamin-related genes revealed the lack of *Cbi*N – a cobalt transporter protein among the *Giardia* positive groups, suggesting an indirect impact on activating cobalamin and discordance at the individual vital amino acid level. In a larger study, children with giardiasis had broad amino acid deficiencies(56). The available data in this study cannot determine whether the lack of the *Cbi*N gene impacts the overall cobalamin availability across the gut microbiome.

Limitations of this study include the lack of statistical power across relatively small samples of children sampled at 3 and 5 years of age. Also, serum levels of cobalamin were unavailable, and metabolomics data could not correlate with microbiome findings or anthropometric measures. Finally, a gap of a two-year timeframe between sample collection may be comparatively long to detect microbiome changes.

## Conclusion

This longitudinal analysis observed a decrease in alpha diversity normalized to *Giardia* DNA (fg/µl) concentration as children increased in age from 3 to 5 years old, revealing the impact on microbiome diversity changes as children age. Differential taxa abundance analysis showed microbiome changes at genus and species levels, which could allow biofilm formation and reduction in potentially beneficial butyrate-producing bacteria. There were also fewer cobalamin synthesis genes among the microbiomes of the children with a higher infectious burden of *Giardia*. These findings may help understand the mechanism of how *Giardia* may lead to growth delays and stunting.

## Data Availability

The data supporting the conclusions of this article will be available in the NCBI BioProject repository upon acceptance.

## Funding

Internal funds from New England Biolabs for DNA sequencing. Research funding was provided by the US Department of Health and Human Services, Health Resources and Services Administration for Baylor College of Medicine Center of Excellence in Health Equity, Training, and Research (Grant No: D34HP31024). Also, the Baylor College of Medicine Junior Faculty Seed Award and BCM Center for Globalization Pilot Project were used for research supplies. Also, Wellcome Trust grant 088862/Z/09/Z.

## Authors’ contributions

RM and AD were responsible for bioinformatics and statistical analysis. RM, AA, AL performed DNA extraction, qPCR, demethylation, and library preparation. EL, CL, ED, BS were involved in the DNA sequencing sample reactions and preliminary analysis. RM, AD, VSH, DGR, BS, PJC wrote the manuscript. All authors read and approved the final manuscript.

## Notes

### Competing Interest Statement

The authors have declared no competing interest.

### Funding Statement

Yes

## References

1. Certad G, Viscogliosi E, Chabé M, Cacciò SM. Pathogenic Mechanisms of Cryptosporidium and Giardia. Trends Parasitol. 2017 Jul 1;33(7):561–76.

2. Halliez MC, Buret AG. Extra-intestinal and long term consequences of Giardia duodenalis infections. World J Gastroenterol WJG. 2013 Dec 21;19(47):8974–85.

3. Beatty JK, Akierman SV, Motta JP, Muise S, Workentine ML, Harrison JJ, et al. Giardia duodenalis induces pathogenic dysbiosis of human intestinal microbiota biofilms. Int J Parasitol. 2017 May 1;47(6):311–26.

4. Huth S von, Thingholm LB, Kofoed PE, Bang C, Rühlemann MC, Franke A, et al. Intestinal protozoan infections shape fecal bacterial microbiota in children from Guinea-Bissau. PLoS Negl Trop Dis. 2021 Mar 3;15(3):e0009232.

5. Riba A, Hassani K, Walker A, Best N van, Zeschwitz D von, Anslinger T, et al. Disturbed gut microbiota and bile homeostasis in Giardia-infected mice contributes to metabolic dysregulation and growth impairment. Sci Transl Med [Internet]. 2020 Oct 14 [cited 2021 Mar 9];12(565). Available from: https://stm.sciencemag.org/content/12/565/eaay7019

6. Toro-Londono MA, Bedoya-Urrego K, Garcia-Montoya GM, Galvan-Diaz AL, Alzate JF. Intestinal parasitic infection alters bacterial gut microbiota in children. PeerJ. 2019 Jan 7;7:e6200.

7. Jacobsen KH, Ribeiro PS, Quist BK, Rydbeck BV. Prevalence of intestinal parasites in young Quichua children in the highlands of rural Ecuador. J Health Popul Nutr. 2007 Dec;25(4):399–405.

8. Lowenstein C, Vasco K, Sarzosa S, Salinas L, Torres A, Perry MJ, et al. Determinants of Childhood Zoonotic Enteric Infections in a Semirural Community of Quito, Ecuador. Am J Trop Med Hyg. 2020 Mar 30;102(6):1269–78.

9. Mejia R, Vicuña Y, Broncano N, Sandoval C, Vaca M, Chico M, et al. A Novel, Multi-Parallel, Real-Time Polymerase Chain Reaction Approach for Eight Gastrointestinal Parasites Provides Improved Diagnostic Capabilities to Resource-Limited At-Risk Populations. Am J Trop Med Hyg. 2013 Jun 5;88(6):1041–7.

10. Bryan PE, Romero M, Sánchez M, Torres G, Gómez W, Restrepo M, et al. Urban versus Rural Prevalence of Intestinal Parasites Using Multi-Parallel qPCR in Colombia. Am J Trop Med Hyg. 2020 Dec 14;

11. Fantinatti M, Lopes-Oliveira LAP, Cascais-Figueredo T, Austriaco-Teixeira P, Verissimo E, Bello AR, et al. Recirculation of Giardia lamblia Assemblage A After Metronidazole Treatment in an Area With Assemblages A, B, and E Sympatric Circulation. Front Microbiol [Internet]. 2020 Oct 22 [cited 2021 Mar 6];11. Available from: https://www.ncbi.nlm.nih.gov/pmc/articles/PMC7642054/

12. Forgie AJ, Drall KM, Bourque SL, Field CJ, Kozyrskyj AL, Willing BP. The impact of maternal and early life malnutrition on health: a diet-microbe perspective. BMC Med [Internet]. 2020 May 12 [cited 2021 Mar 26];18. Available from: https://www.ncbi.nlm.nih.gov/pmc/articles/PMC7216331/

13. Green R, Allen LH, Bjørke-Monsen AL, Brito A, Guéant JL, Miller JW, et al. Vitamin B 12 deficiency. Nat Rev Dis Primer. 2017 Jun 29;3(1):1–20.

14. Langan RC, Goodbred AJ. Vitamin B12 Deficiency: Recognition and Management. Am Fam Physician. 2017 Sep 15;96(6):384–9.

15. Goraya JS. Vitamin B12 deficiency in Indian infants and children. Paediatr Int Child Health. 2020 May;40(2):75–7.

16. Handbook of Vitamins [Internet]. CRC Press; 2013 [cited 2021 Mar 9]. Available from: https://www.taylorfrancis.com/books/handbook-vitamins-janos-zempleni-john-suttie-jesse-gregory-iii-patrick-stover/10.1201/b15413

17. Mach N, Clark A. Micronutrient Deficiencies and the Human Gut Microbiota. Trends Microbiol. 2017 Aug 1;25(8):607–10.

18. Degnan PH, Taga ME, Goodman AL. Vitamin B 12 as a Modulator of Gut Microbial Ecology. Cell Metab. 2014 Nov;20(5):769–78.

19. Boran P, Baris HE, Kepenekli E, Erzik C, Soysal A, Dinh DM. The impact of vitamin B12 deficiency on infant gut microbiota. Eur J Pediatr. 2020 Mar 1;179(3):385–93.

20. Rizowy GM, Poloni S, Colonetti K, Donis KC, Dobbler PT, Leistner-Segal S, et al. Is the gut microbiota dysbiotic in patients with classical homocystinuria? Biochimie. 2020 Jun;173:3– 11.

21. Mejia R, Damania A, Jeun R, Bryan PE, Vargas P, Juarez M, et al. Impact of intestinal parasites on microbiota and cobalamin gene sequences: a pilot study. Parasit Vectors. 2020 Apr 19;13(1):200.

22. Von Huth S, Thingholm LB, Kofoed PE, Bang C, Rühlemann MC, Franke A, et al. Intestinal protozoan infections shape fecal bacterial microbiota in children from Guinea-Bissau. Bartelt LA, editor. PLoS Negl Trop Dis. 2021 Mar 3;15(3):e0009232.

23. Cooper PJ, Chico ME, Platts-Mills TA, Rodrigues LC, Strachan DP, Barreto ML. Cohort Profile: The Ecuador Life (ECUAVIDA) study in Esmeraldas Province, Ecuador. Int J Epidemiol. 2015 Oct;44(5):1517–27.

24. Cimino RO, Jeun R, Juarez M, Cajal PS, Vargas P, Echazú A, et al. Identification of human intestinal parasites affecting an asymptomatic peri-urban Argentinian population using multi-parallel quantitative real-time polymerase chain reaction. Parasit Vectors. 2015 Jul 17;8:380.

25. Bushnell B. BBMap: A Fast, Accurate, Splice-Aware Aligner [Internet]. Lawrence Berkeley National Lab. (LBNL), Berkeley, CA (United States); 2014 Mar [cited 2021 Apr 2]. Report No.: LBNL-7065E. Available from: https://www.osti.gov/biblio/1241166-bbmap-fast-accurate-splice-aware-aligner

26. Chen S, Zhou Y, Chen Y, Gu J. fastp: an ultra-fast all-in-one FASTQ preprocessor. Bioinformatics. 2018 Sep 1;34(17):i884–90.

27. Ewels P, Magnusson M, Lundin S, Käller M. MultiQC: summarize analysis results for multiple tools and samples in a single report. Bioinformatics. 2016 Oct 1;32(19):3047–8.

28. O’Leary NA, Wright MW, Brister JR, Ciufo S, Haddad D, McVeigh R, et al. Reference sequence (RefSeq) database at NCBI: current status, taxonomic expansion, and functional annotation. Nucleic Acids Res. 2016 Jan 4;44(D1):D733–745.

29. Buchfink B, Xie C, Huson DH. Fast and sensitive protein alignment using DIAMOND. Nat Methods. 2015 Jan;12(1):59–60.

30. Blum M, Chang HY, Chuguransky S, Grego T, Kandasaamy S, Mitchell A, et al. The InterPro protein families and domains database: 20 years on. Nucleic Acids Res. 2021 Jan 8;49(D1):D344–54.

31. Huson DH, Beier S, Flade I, Górska A, El-Hadidi M, Mitra S, et al. MEGAN Community Edition - Interactive Exploration and Analysis of Large-Scale Microbiome Sequencing Data. PLOS Comput Biol. 2016 Jun 21;12(6):e1004957.

32. Parks DH, Tyson GW, Hugenholtz P, Beiko RG. STAMP: statistical analysis of taxonomic and functional profiles. Bioinforma Oxf Engl. 2014 Nov 1;30(21):3123–4.

33. McMurdie PJ, Holmes S. phyloseq: An R Package for Reproducible Interactive Analysis and Graphics of Microbiome Census Data. PLOS ONE. 2013 Apr 22;8(4):e61217.

34. R Core Team. R: A Language and Environment for Statistical Computing [Internet]. Vienna, Austria: R Foundation for Statistical Computing; 2020. Available from: https://www.R-project.org/

35. Wickham H, François R, Henry L, Müller K. dplyr: A Grammar of Data Manipulation [Internet]. 2021. Available from: https://CRAN.R-project.org/package=dplyr

36. Segata N, Waldron L, Ballarini A, Narasimhan V, Jousson O, Huttenhower C. Metagenomic microbial community profiling using unique clade-specific marker genes. Nat Methods. 2012 Aug;9(8):811–4.

37. Oksanen J, Blanchet FG, Friendly M, Kindt R, Legendre P, McGlinn D, et al. vegan: Community Ecology Package [Internet]. 2020. Available from: https://CRAN.R-project.org/package=vegan

38. Mallick H, Rahnavard A, McIver LJ, Ma S, Zhang Y, Nguyen LH, et al. Multivariable association discovery in population-scale meta-omics studies. PLOS Comput Biol. 2021 Nov 16;17(11):e1009442.

39. Martin BD, Witten D, Willis AD. Modeling microbial abundances and dysbiosis with beta-binomial regression. Ann Appl Stat. 2020 Mar;14(1):94–115.

40. Kuznetsova A, Brockhoff PB, Christensen RHB. lmerTest Package: Tests in Linear Mixed Effects Models. J Stat Softw. 2017;82(13):1–26.

41. Wickham H. ggplot2: elegant graphics for data analysis. Second edition. Cham: Springer; 2016. 260 p. (Use R!).

42. Dunn N, Juergens AL. Giardiasis. In: StatPearls [Internet]. Treasure Island (FL): StatPearls Publishing; 2021 [cited 2021 Apr 16]. Available from: http://www.ncbi.nlm.nih.gov/books/NBK513239/

43. Roswall J, Olsson LM, Kovatcheva-Datchary P, Nilsson S, Tremaroli V, Simon MC, et al. Developmental trajectory of the healthy human gut microbiota during the first 5 years of life. Cell Host Microbe [Internet]. 2021 Mar 31 [cited 2021 Apr 24];0(0). Available from: https://www.cell.com/cell-host-microbe/abstract/S1931-3128(21)00100-1

44. Barash NR, Maloney JG, Singer SM, Dawson SC. Giardia Alters Commensal Microbial Diversity throughout the Murine Gut. Infect Immun [Internet]. 2017 May 23 [cited 2021 Apr 17];85(6). Available from: https://www.ncbi.nlm.nih.gov/pmc/articles/PMC5442636/

45. McGregor BA, Razmjou E, Hooshyar H, Seeger DR, Golovko SA, Golovko MY, et al. A shotgun metagenomic analysis of the fecal microbiome in humans infected with Giardia duodenalis. Parasit Vectors. 2023 Jul 18;16(1):239.

46. Toro-Londono MA, Bedoya-Urrego K, Garcia-Montoya GM, Galvan-Diaz AL, Alzate JF. Intestinal parasitic infection alters bacterial gut microbiota in children. PeerJ. 2019 Jan 7;7:e6200.

47. Mejia R, Damania A, Jeun R, Bryan PE, Vargas P, Juarez M, et al. Impact of intestinal parasites on microbiota and cobalamin gene sequences: a pilot study. Parasit Vectors. 2020 Dec;13(1):200.

48. Donaldson GP, Lee SM, Mazmanian SK. Gut biogeography of the bacterial microbiota. Nat Rev Microbiol. 2016 Jan;14(1):20–32.

49. Martinez-Guryn K, Leone V, Chang EB. Regional Diversity of the Gastrointestinal Microbiome. Cell Host Microbe. 2019 Sep 11;26(3):314–24.

50. Motta JP, Wallace JL, Buret AG, Deraison C, Vergnolle N. Gastrointestinal biofilms in health and disease. Nat Rev Gastroenterol Hepatol. 2021 Jan 28;1–21.

51. Tytgat HLP, Nobrega FL, van der Oost J, de Vos WM. Bowel Biofilms: Tipping Points between a Healthy and Compromised Gut? Trends Microbiol. 2019 Jan 1;27(1):17–25.

52. Fekete E, Allain T, Siddiq A, Sosnowski O, Buret AG. Giardia spp. and the Gut Microbiota: Dangerous Liaisons. Front Microbiol [Internet]. 2021 Jan 12 [cited 2021 Feb 23];11. Available from: https://www.ncbi.nlm.nih.gov/pmc/articles/PMC7835142/

53. Scheiman J, Luber JM, Chavkin TA, MacDonald T, Tung A, Pham LD, et al. Meta-omics analysis of elite athletes identifies a performance-enhancing microbe that functions via lactate metabolism. Nat Med. 2019 Jul;25(7):1104–9.

54. Rowley CA, Kendall MM. To B12 or not to B12: Five questions on the role of cobalamin in host-microbial interactions. PLOS Pathog. 2019 Jan 3;15(1):e1007479.

55. Wexler AG, Schofield WB, Degnan PH, Folta-Stogniew E, Barry NA, Goodman AL. Human gut Bacteroides capture vitamin B12 via cell surface-exposed lipoproteins. eLife. 2018 Sep 18;7:e37138.

56. Giallourou N, Arnold J, McQuade ETR, Awoniyi M, Becket RVT, Walsh K, et al. Giardia hinders growth by disrupting nutrient metabolism independent of inflammatory enteropathy. Nat Commun. 2023 May 18;14(1):2840.

